# Ranitidine use, N-Nitrosodimethylamine (NDMA) production and variations in cancer diagnoses

**DOI:** 10.1101/2021.01.27.21250656

**Authors:** Lior Z. Braunstein, Elizabeth D. Kantor, Kelli O’Connell, Amber Hudspeth, Kaury Kucera, Qian Wu, David Y. Light

## Abstract

**Introduction:** Ranitidine is a member of the H_2_-blocker class of medications, commonly used in the treatment gastroesophageal reflux and peptic ulcer disease. Recent laboratory analyses have demonstrated that ranitidine may yield the probable carcinogen, N-Nitrosodimethylamine (NDMA). Here, we characterized the kinetics of NDMA production from ranitidine under various simulated gastric conditions. Moreover, we conducted a cross-sectional epidemiologic analysis to evaluate potential associations between ranitidine use and various cancer presentations.

**Methods:** The formation of NDMA in the presence of ranitidine was studied in a series of experiments conducted in simulated gastric fluid (SGF), while varying pH, nitrite concentration, and time of reaction. Chemical analyses of NDMA yield were performed by liquid chromatography-high resolution mass spectrometry per FDA guidelines on the detection of NDMA from ranitidine drug products.

To evaluate potential associations between ranitidine use and cancer diagnoses, we conducted a cross-sectional analysis among a population of oncology patients, following institutional review board approval. Analyses were limited to those reporting use of ranitidine or an active comparator (i.e. proton-pump inhibitor [PPI] or other H_2_-blocker) to partially mitigate potential confounding. Multivariable logistic regression was employed to identify associations between ranitidine and certain cancers, adjusted for age, sex, race, ethnicity, and body-mass index.

**Results:** Increasing sodium nitrite concentrations in SGF at pH 1.2 were associated with increasing NDMA yield. At a pH of 2.5, previously identified as the optimal reaction condition, samples collected at incubation times of 1, 2 and 4 hours (approximating typical gastric emptying conditions) showed a continued increase in NDMA over time. Notably, the reaction trajectory suggests NDMA may have continued to form beyond the last observed timepoint of 4 hours.

On cross-sectional analysis of an oncology population, use of ranitidine (versus active comparators) was associated with increased odds of presenting with cancers of the breast (OR=1.58; 95%CI: 1.23-2.01) as compared to presentation with another cancer. Similar positive associations were observed for cancers of the thyroid (OR=1.89; 95%CI: 1.18-2.92), bladder (OR=1.58; 95%CI: 1.10-2.21), and prostate (OR=1.80; 95%CI: 1.34-2.39). Conversely, ranitidine was inversely associated with cancers of the colorectum (OR=0.48; 95%CI: 0.30-0.74) and brain (OR=0.56; 95%CI: 0.33-0.89).

**Conclusion:** Under simulated gastric conditions, ranitidine yields increasing amounts of NDMA over time (up to, and likely beyond 4 hours) and with increasing concentrations of sodium nitrite. In addition, among a cohort of cancer patients reporting use of H_2_-blockers or PPIs at the time of diagnosis, we found an association between ranitidine use and cancers of the breast, thyroid, bladder and prostate. We further observed unexplained inverse associations with cancers of the brain and colorectum. These associations reflect the odds of presenting with different cancers exclusively among an oncology population, and do not directly represent risk in a general population (as there were no cancer-free individuals in this study). Moreover, these exploratory analyses are to be viewed as hypothesis generating and should prompt analyses of larger cohorts with longer follow-up.

## Introduction

Ranitidine (brand name “Zantac”) is a small molecule drug in the H_2_-receptor antagonist class of agents (i.e. “H_2_-blockers”). First marketed in the United States in 1983 as an inhibitor of gastric acid secretion for the treatment of gastroesophageal reflux and peptic ulcer disease, ranitidine was thereafter added to the World Health Organization’s List of Essential Medicines.^1^

The ranitidine molecule is a tertiary amine, a class of drugs that have been posited to yield carcinogenic nitrosamine species under physiologic conditions.^2^ To that end, recent analyses show that commercially-available ranitidine degrades to form high levels of N-Nitrosodimethylamine (NDMA), a probable human carcinogen and a potent carcinogen in animal models.^3,4^ These findings recently prompted the European Medicines Agency^5^ and US Food and Drug Administration to request that sales of all ranitidine products be halted.^6^

Despite the established link between ranitidine and NDMA, the association between ranitidine and cancer in humans remains unclear. Few epidemiologic studies have evaluated the potential link between ranitidine and cancer, with some retrospective analyses suggesting elevated risks of breast, esophageal and gastric cancers, ^7,8^ and others demonstrating no excess cancer risk.^9^

Herein, we sought to further characterize the chemistry of NDMA formation from ranitidine under simulated gastric conditions. Moreover, in light of the potential for ranitidine to confer carcinogenic risk via NDMA exposure, we evaluated the association between ranitidine use and cancer risk in a cohort of oncology patients who reported taking ranitidine or other H_2_-blockers/proton pump inhibitors (PPIs) around the time of their cancer diagnoses.

## Methods

### Chemical analyses

To characterize the formation and fate of NDMA under physiologically relevant conditions in the presence of ranitidine, a series of experiments was conducted in simulated gastric fluid (SGF), varying pH, nitrite concentration, and time of reaction. The chemical analyses of NDMA were performed on a liquid chromatography-high resolution mass spectrometer.^10^

### Equipment, Supplies, and Chemicals

SCIEX EXIONLC AD (LC) coupled with an X500R time of flight high resolution mass spectrometer (HRMS) was purchased from SCIEX (Framingham, MA). An Agilent Infinity Poroshell 120 EC-C18 HPLC column (2.7 µm, 4.6 × 100 mm) was purchased from Agilent (Santa Clara, CA). Certified reference materials (CRM) of N-Nitrosodimethylamine (NDMA) (≥ 99.2 %) were purchased from Sigma-Aldrich (St. Louis, MO). Isotopic labeled NDMA standard ^13^C_2_-D_6_-NDMA was purchased from Cambridge Isotope Laboratories (Tewksbury, MA). All other chemicals and reagents were ACS grade from Sigma-Aldrich. Ranitidine prescription 150 mg and “cool mint” flavored branded ranitidine 150 mg (“Zantac”) were compared. All tablets of each of the two varieties were obtained from the same lot, and lots were sampled well within their labelled expiration dates.

### Experiments

SGF (2g/L sodium chloride in water) was prepared at various pH and nitrite concentrations. Whereas we previously reported that NDMA formation at pH 1.2 to 3.5 was higher than pH 4.5 to 5.5 [Braunstein LZ, et al. *in press]*, herein we sought to further elucidate the kinetics of NDMA formation at variable nitrite concentrations in 100 mL SGF containing 50, 25, 10, and 2.5 mM nitrite at pH 1.2 and 2.5, the previously determined peak reaction condition. Ranitidine prescription 150 mg and “cool mint” flavored branded ranitidine 150 mg were added to SGF. All experiments were conducted at 37 °C incubation, and samples were taken at 1, 2, and 4-hour time points. At the designated time points, one milliliter (1 mL) of incubated SGF was transferred, centrifuged, and filtered by a 0.2 µm nylon filter into an HPLC vial. A known amount of ^13^C_2_-D_6_-NDMA was spiked into the sample targeting the final internal standard concentration of 40 ng/mL. The prescription ranitidine and branded “cool mint” ranitidine tablets (n=5 each) were also directly diluted in methanol (Sigma-Aldrich LC-MS grade) and run by LC-HRMS as control samples to determine any background of NDMA contamination. The average NDMA background was subtracted from experimental data.

### Instrumental Analysis

NDMA concentration in the SGF solution was determined by a method modified from the FDA recommended liquid chromatography – high resolution mass spectrometry (LC-HRMS) method for determination of nitrosamines in ranitidine drug substance and drug products, and was able to eliminate interference from a heavy isotope of dimethylformamide.^10^ Briefly, chromatographic separation started at a gradient of 98% of mobile phase A (0.1% formic acid in water) and 2% of mobile phase B (0.1% formic acid in methanol) and was held for 3.5 minutes. Mobile phase B ramped up to 98% at 9.5 minutes and was held for 2.4 minutes, followed by B ramped down to 2% and held to 15 minutes. Total HPLC flow rate was 0.6 mL/min. NDMA elutes at 4.44 minutes. Atmospheric pressure chemical ionization mode (APCI+) was selected to ionize NDMA and its isotopic labeled internal standard. Mass identification for NDMA was done by detecting the accurate mass of [M+1]^+^ in MRMHR acquisition mode for NDMA 75.0553>75.0553, and ^13^C_2_-D_6_-NDMA 83.0997>83.0997, respectively. The mass accuracy was set at 5 parts per million (ppm) and mass resolution was greater than 31,000 for mass 176.09134 Da in the APCI+ calibrant solution.

### Quality Assurance and Quality Control

The quadratic calibration curve was established by a nine-point calibration ranging from 0.25 to 200 ng/mL containing the same internal standard concentration as samples. Calibration was accepted if the r^2^ was ≥ 0.99. The limit of quantification (LOQ) was defined as the lowest acceptable calibration point. A calibration standard was acceptable if the calculated concentration was within ± 20% of its theoretical concentration, except for the lowest acceptable concentration which was ± 30%. The lowest calibration point was a minimum signal to noise ratio of 10. Concentrations of NDMA in samples were quantified by the internal standard method. The LOQ was 0.25 ng/mL for NDMA and was equivalent to 25 ng in each sample incubation. When NDMA concentration in sample extracts exceeded the calibration range, sample extracts were diluted with 3:1 water:methanol solution containing internal standard for re-analysis. Low, medium, and high QC samples were prepared following the same procedure of sample preparation, and all QCs are within ± 20% of their theoretical concentrations.

### Cross-sectional Epidemiologic Analysis

To assess potential epidemiologic implications of ranitidine exposure, following institutional review board approval we conducted a cross-sectional analysis of ranitidine use in relation to cancer diagnosis among patients newly presenting to Memorial Sloan Kettering Cancer Center with localized invasive or *in situ* disease between 2013 and 2019. Patients who reported use of an H_2_-blocker or proton-pump inhibitor (PPI) about the time of initial cancer diagnosis were included. Multivariable logistic regression was employed to identify associations between ranitidine (as opposed to other H_2_-blockers and PPIs) and certain cancers, adjusted for age, sex, race, ethnicity, and body-mass index (BMI). Age was measured as a grouped linear variable with the following groups: <50, 50-<60, 60-<70, 70-<80, and 80+ years. Race was categorized as White, Black, and Asian/Other/Unknown. Ethnicity was categorized as Non-Hispanic, Hispanic, and Unknown. BMI was a categorical variable with categories <25 kg/m^2^, 25-<30 kg/m^2^, 30+ kg/m^2^, and missing. Models for breast, ovarian, and endometrial cancers were restricted to women and therefore were not adjusted for sex. Similarly, the model for prostate cancer was restricted to men and was not adjusted for sex. To address potential for confounding, analyses were limited to persons reporting use of ranitidine or an active comparator (i.e. PPIs or other H_2_-blockers). Of note, resultant odds ratios represent the odds of presenting with the specified cancer as compared to presenting with other cancers (not as compared to having no cancer, since no such cancer-free patients were included in the study).

## Results

To characterize the formation of NDMA under various reactive conditions, we optimized the FDA-recommended LC-HRMS assay for detection of NDMA from ranitidine products. As described above, we tested prescription ranitidine 150mg and branded “cool-mint” ranitidine 150mg under each reactive condition (see methods). Notably, prescription ranitidine exhibited very low endogenous NDMA (7 ng/tablet ± 11%), while the branded “cool mint” ranitidine yielded a substantial background of NDMA (4590 ng/tablet ± 10%). Background levels of NDMA were subtracted from experimental analytic results to better represent only that quantity of NDMA formed during the described incubations.

At pH 1.2, increasing sodium nitrite concentration was associated with increasing NDMA detection (Figure 1A). Samples collected at incubation times of 1, 2 and 4 hours (approximating typical gastric emptying durations) at pH 2.5, previously identified as being both physiologically relevant and the optimal reaction pH, showed a continued increase in NDMA over time (Figure 1B). Trends in the kinetics of NDMA formation were consistent across the generic prescription and branded over-the counter formulations studied.

**Figure 1:**
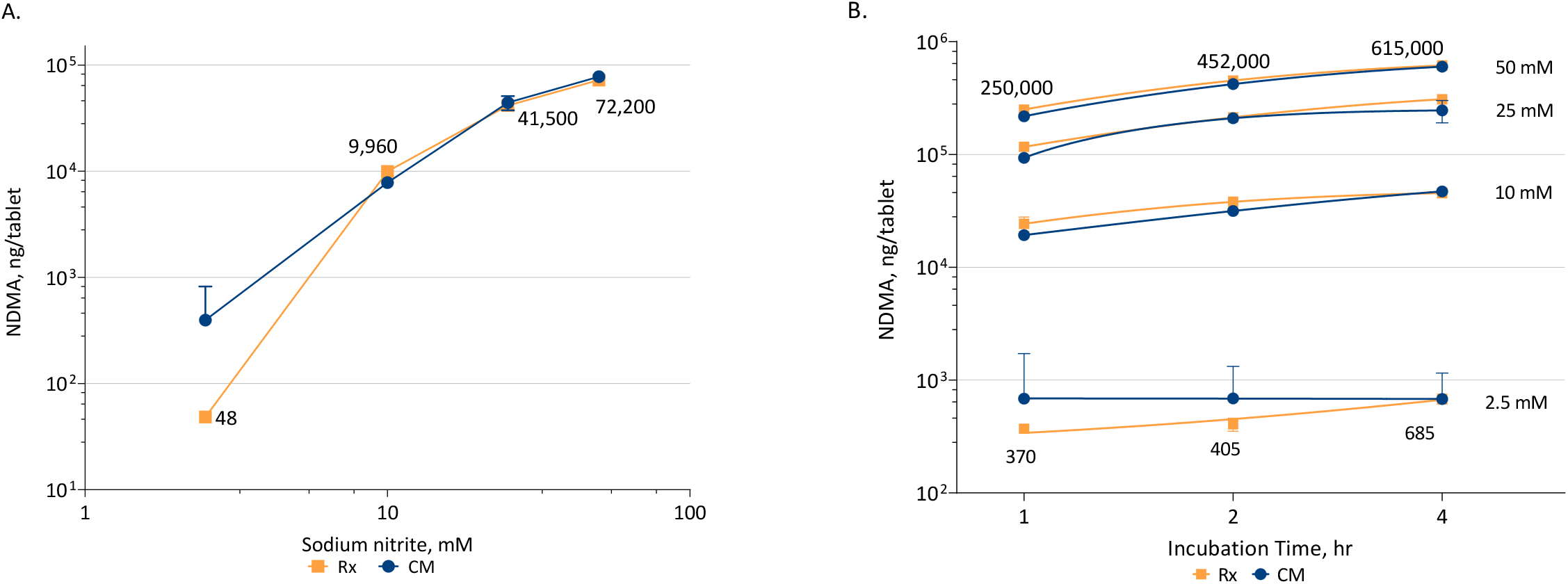
NDMA formation from branded ranitidine 150 mg “Cool Mint” (CM) and prescription ranitidine 150mg (Rx). A) Incubation at pH 1.2 with varying concentrations of sodium nitrite (NaNO_2_) for a typical gastric-emptying time of 2 hours. B) Incubation at pH 2.5 with varying concentrations of NaNO_2_ (2.5, 10, 25, and 50mM) and evaluated over a range of incubation times (1, 2, and 4 hours). Error bars represent the standard deviation of data from triplicate reactions. Due to substantial endogenous NDMA in the CM tablets, conditions resulting in relatively low NDMA formation in CM tablets display considerable quantification variability. Numerical labels of NDMA formation amounts are for Rx tablets only.

To evaluate potential epidemiologic implications of these findings, we analyzed the association between ranitidine use and various cancer presentations in a population of oncology patients who reported use of either ranitidine or another H_2_-blocker/PPI at the time of diagnosis (mean age was 63.9 years; 50.0% were male; table 1). At exposure assessment, 8.4% (n=871) reported ranitidine use, while the remainder used an active comparator (n=9,476).

**Table 1.** Sociodemographic Characteristics of Study Population, by Exposure Status

|  | **Total**<br>**(N=10347)** | **Ranitidine Only**<br>**(N=871)** | **Active Comparator**<br>**(other H2 blocker/PPI)**<br>**(N=9476)** |
| --- | --- | --- | --- |
| **Age (years)** |  |  |  |
| <50 | 1419 (13.7%) | 123 (14.1%) | 1296 (13.7%) |
| 50-<60 | 2024 (19.6%) | 180 (20.7%) | 1844 (19.5%) |
| 60-<70 | 3180 (30.7%) | 276 (31.7%) | 2904 (30.6%) |
| 70-<80 | 2725 (26.3%) | 223 (25.6%) | 2502 (26.4%) |
| 80+ | 999 (9.7%) | 69 (7.9%) | 930 (9.8%) |
| **Sex** |  |  |  |
| Male | 5172 (50.0%) | 386 (44.3%) | 4786 (50.5%) |
| Female | 5175 (50.0%) | 485 (55.7%) | 4690 (49.5%) |
| **Race** |  |  |  |
| White | 8634 (83.4%) | 741 (85.1%) | 7893 (83.3%) |
| Black | 477 (4.6%) | 34 (3.9%) | 443 (4.7%) |
| Asian | 495 (4.8%) | 40 (4.6%) | 455 (4.8%) |
| Other/Unknown | 741 (7.2%) | 56 (6.4%) | 685 (7.2%) |
| **Ethnicity** |  |  |  |
| Non-Hispanic | 9288 (89.8%) | 780 (89.6%) | 8508 (89.8%) |
| Hispanic | 559 (5.4%) | 48 (5.5%) | 511 (5.4%) |
| Unknown | 500 (4.8%) | 43 (4.9%) | 457 (4.8%) |
| **BMI (kg/m<sup>2</sup>)** |  |  |  |
| <25 | 2582 (25.0%) | 256 (29.4%) | 2326 (24.5%) |
| 25-<30 | 3044 (29.4%) | 243 (27.9%) | 2801 (29.6%) |
| 30+ | 2805 (27.1%) | 204 (23.4%) | 2601 (27.4%) |
| Missing | 1916 (18.5%) | 168 (19.3%) | 1748 (18.4%) |

Use of ranitidine (versus active comparators), was associated with a significant increase in the adjusted odds ratio (OR) of presenting with cancers of the breast (OR=1.58; 95%CI: 1.23-2.01). thyroid (OR=1.89; 95%CI: 1.18-2.92), bladder (OR=1.58; 95%CI: 1.10-2.21), and prostate (OR=1.80; 95%CI: 1.34-2.39 (Table 2; Figure 2) as compared to presentations with another cancer. Inverse associations were observed for colorectal cancer (OR=0.48; 95%CI: 0.30-0.74) and brain malignancies (OR=0.56; 95%CI: 0.33-0.89) (Figure 2). No association was observed for cancers of the lung, kidney, skin (melanoma), blood (leukemias and lymphomas), ovary or endometrium. In sensitivity analyses excluding *in situ* cases, interpretations remained unchanged with the exception of bladder cancer which attenuated and became non-significant (OR=1.41; 95%CI:0.91-2.07) (Table 3).

**Table 2.**
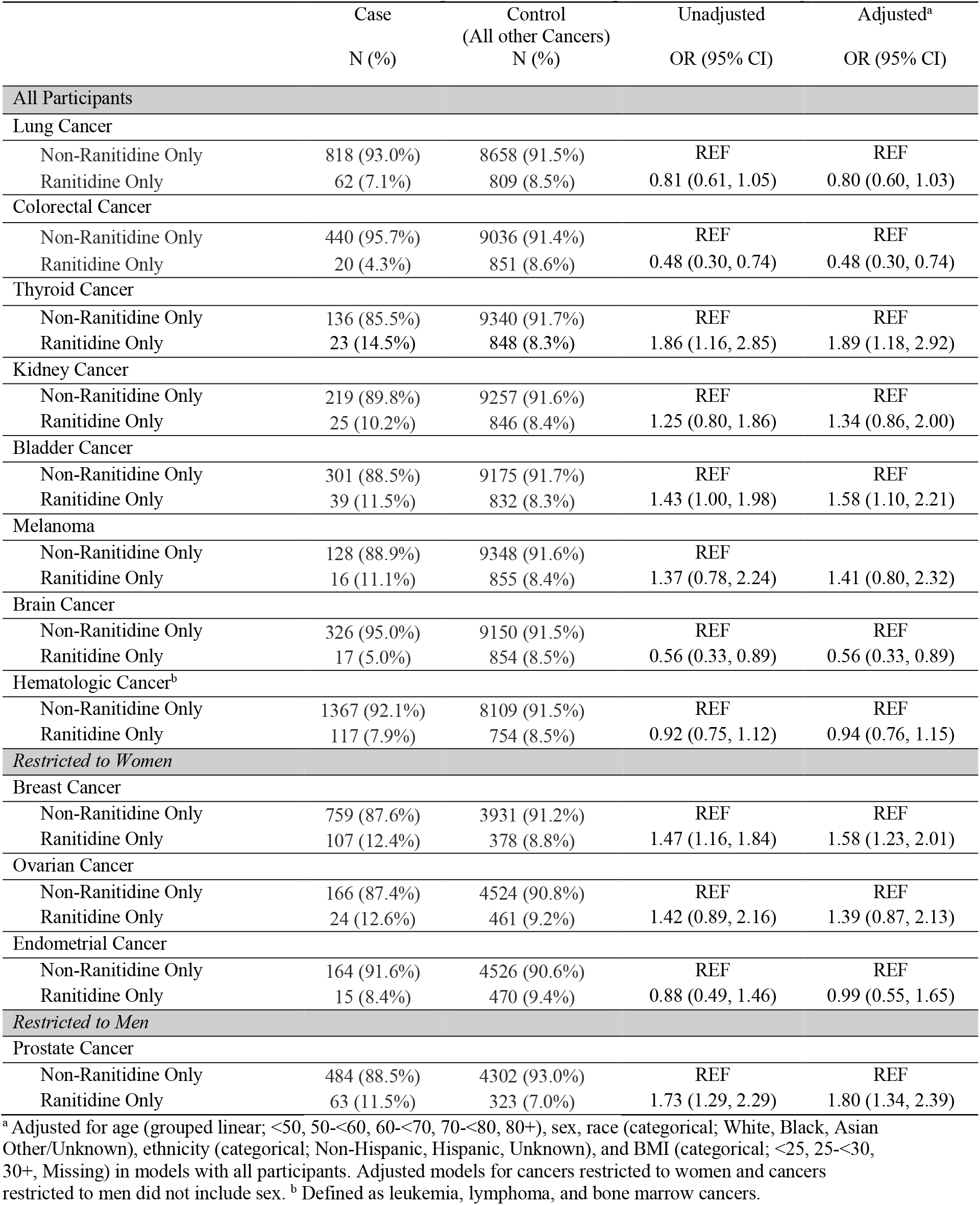
Associations between Ranitidine vs Active Comparator Use and Site-Specific Cancer (including in situ)

**Table 3.**
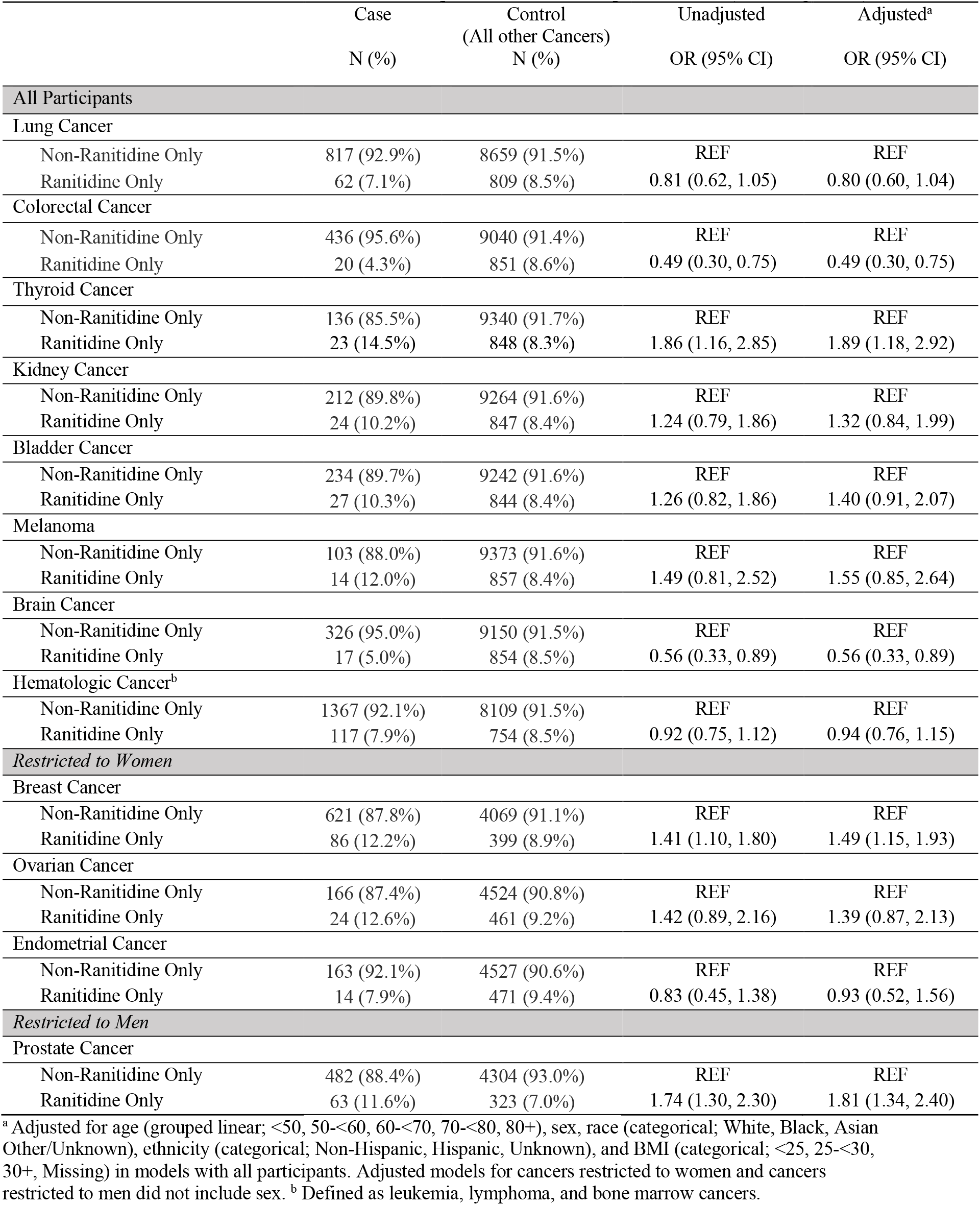
Associations between Ranitidine vs Active Comparator Use and Site-Specific Cancer (excluding in situ)

|  | Case<br>N (%) | Control<br>(All other Cancers)<br>N (%) | Unadjusted<br>OR (95% CI) | Adjusted<sup>a</sup><br>OR (95% CI) |
| --- | --- | --- | --- | --- |
| **All Participants** |  |  |  |  |
| **Lung Cancer** |  |  |  |  |
| Non-Ranitidine Only | 817 (92.9%) | 8659 (91.5%) | REF | REF |
| Ranitidine Only | 62 (7.1%) | 809 (8.5%) | 0.81 (0.62, 1.05) | 0.80 (0.60, 1.04) |
| **Colorectal Cancer** |  |  |  |  |
| Non-Ranitidine Only | 436 (95.6%) | 9040 (91.4%) | REF | REF |
| Ranitidine Only | 20 (4.3%) | 851 (8.6%) | 0.49 (0.30, 0.75) | 0.49 (0.30, 0.75) |
| **Thyroid Cancer** |  |  |  |  |
| Non-Ranitidine Only | 136 (85.5%) | 9340 (91.7%) | REF | REF |
| Ranitidine Only | 23 (14.5%) | 848 (8.3%) | 1.86 (1.16, 2.85) | 1.89 (1.18, 2.92) |
| **Kidney Cancer** |  |  |  |  |
| Non-Ranitidine Only | 212 (89.8%) | 9264 (91.6%) | REF | REF |
| Ranitidine Only | 24 (10.2%) | 847 (8.4%) | 1.24 (0.79, 1.86) | 1.32 (0.84, 1.99) |
| **Bladder Cancer** |  |  |  |  |
| Non-Ranitidine Only | 234 (89.7%) | 9242 (91.6%) | REF | REF |
| Ranitidine Only | 27 (10.3%) | 844 (8.4%) | 1.26 (0.82, 1.86) | 1.40 (0.91, 2.07) |
| **Melanoma** |  |  |  |  |
| Non-Ranitidine Only | 103 (88.0%) | 9373 (91.6%) | REF | REF |
| Ranitidine Only | 14 (12.0%) | 857 (8.4%) | 1.49 (0.81, 2.52) | 1.55 (0.85, 2.64) |
| **Brain Cancer** |  |  |  |  |
| Non-Ranitidine Only | 326 (95.0%) | 9150 (91.5%) | REF | REF |
| Ranitidine Only | 17 (5.0%) | 854 (8.5%) | 0.56 (0.33, 0.89) | 0.56 (0.33, 0.89) |
| **Hematologic Cancer<sup>b</sup>** |  |  |  |  |
| Non-Ranitidine Only | 1367 (92.1%) | 8109 (91.5%) | REF | REF |
| Ranitidine Only | 117 (7.9%) | 754 (8.5%) | 0.92 (0.75, 1.12) | 0.94 (0.76, 1.15) |
| **Restricted to Women** |  |  |  |  |
| **Breast Cancer** |  |  |  |  |
| Non-Ranitidine Only | 621 (87.8%) | 4069 (91.1%) | REF | REF |
| Ranitidine Only | 86 (12.2%) | 399 (8.9%) | 1.41 (1.10, 1.80) | 1.49 (1.15, 1.93) |
| **Ovarian Cancer** |  |  |  |  |
| Non-Ranitidine Only | 166 (87.4%) | 4524 (90.8%) | REF | REF |
| Ranitidine Only | 24 (12.6%) | 461 (9.2%) | 1.42 (0.89, 2.16) | 1.39 (0.87, 2.13) |
| **Endometrial Cancer** |  |  |  |  |
| Non-Ranitidine Only | 163 (92.1%) | 4527 (90.6%) | REF | REF |
| Ranitidine Only | 14 (7.9%) | 471 (9.4%) | 0.83 (0.45, 1.38) | 0.93 (0.52, 1.56) |
| **Restricted to Men** |  |  |  |  |
| **Prostate Cancer** |  |  |  |  |
| Non-Ranitidine Only | 482 (88.4%) | 4304 (93.0%) | REF | REF |
| Ranitidine Only | 63 (11.6%) | 323 (7.0%) | 1.74 (1.30, 2.30) | 1.81 (1.34, 2.40) |

<sup>a</sup> Adjusted for age (grouped linear; <50, 50-<60, 60-<70, 70-<80, 80+), sex, race (categorical; White, Black, Asian Other/Unknown), ethnicity (categorical; Non-Hispanic, Hispanic, Unknown), and BMI (categorical; <25, 25-<30, 30+, Missing) in models with all participants. Adjusted models for cancers restricted to women and cancers restricted to men did not include sex. <sup>b</sup> Defined as leukemia, lymphoma, and bone marrow cancers.

**Figure 2:**
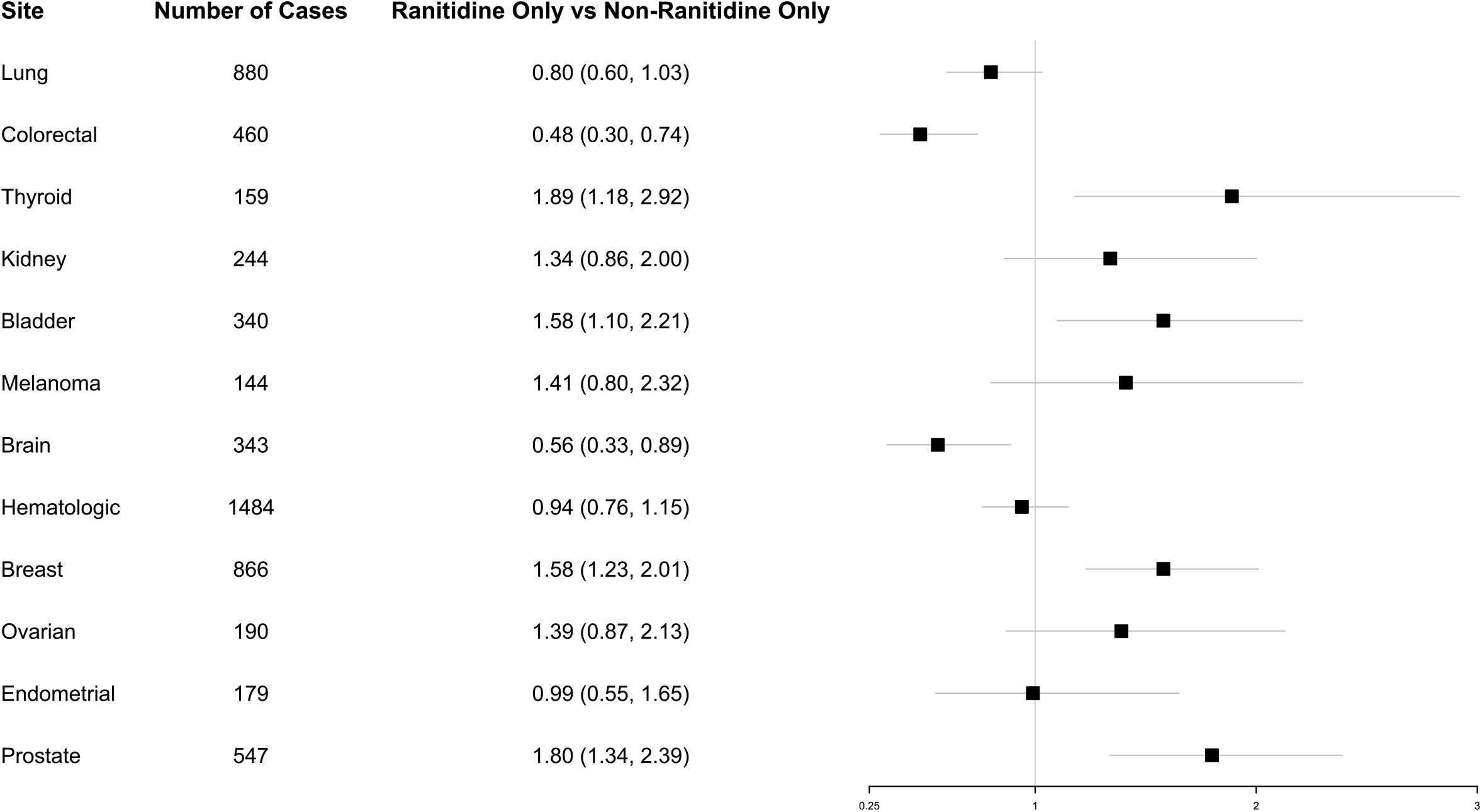
Adjusted odds ratios of presenting cancer diagnoses in an oncology population among those receiving ranitidine versus other H_2_-blockers or proton-pump inhibitors.

## Discussion

In this study, we build on prior work to further characterize conditions under which ranitidine may yield NDMA. We further expand on these findings to report the association between ranitidine use and the odds of presenting with one of several given cancers (among an oncology population). Despite the large cohort size, power was limited for analyses of less-common cancers of interest and these findings merit follow-up in a large, well-powered study.

Since its development nearly four decades ago, ranitidine has rapidly become among the most common agents for treating gastroesophageal reflux and peptic ulcer disease.^11^ Initial studies raised concern that the entire class of H_2_-blockers might yield potential carcinogenic derivatives (i.e. nitroso compounds) due to resultant physiologic alterations in the gastric environment.^12,13^ For example, in a study of 351 subjects, gastric juice contained elevated levels of N-Nitrosamines (including NDMA) in patients given H_2_-blockers, though levels appeared to be lower than established toxic thresholds.^14^

More recently, several groups have specifically focused on the potential for ranitidine, a tertiary amine, to directly form higher levels of NDMA than previously appreciated.^3,4^ While NDMA is a ubiquitous nitrosamine typically found in smoked fish and cured meats,^15^ laboratory studies have shown it to be a potent *in vitro* mutagen via alkylation of DNA.^16,17^ It is also an established carcinogen *in vivo*, yielding cancers of the liver, kidney, and bladder, among others, in animal models.^18-21^ This body of evidence prompted the WHO International Agency for Research on Cancer (IARC) to classify NDMA as probably carcinogenic to humans.^22^ The US Food and Drug Administration has similarly advised that NDMA intake not exceed 96ng/day from drug products.^23^

Recent studies, however, indicate that ranitidine intake may yield NDMA exposure in excess of this established threshold. Filings by an independent pharmacy/analytical laboratory garnered significant public attention upon first revealing that ranitidine tablets generated elevated NDMA levels under simulated gastric conditions.^3^ A separate third-party laboratory further escalated these concerns by identifying excessive NDMA levels in ranitidine drug product that was merely exposed to elevated temperatures.^4^ While the quantity of NDMA per ranitidine tablet has varied over 5 orders of magnitude as a function of the reported assay conditions in each of these studies, none has uniformly exonerated the ranitidine supply. These findings, subsequently confirmed by regulators, have since prompted the US Food and Drug Administration to request that all ranitidine drug products be withdrawn from the US market, that marketing of ranitidine cease, and that market withdrawal plans be submitted by each firm to the government.^24^

Whereas our prior work identified increasing nitrite and pH of 2.5 as optimal reaction conditions for NDMA production from ranitidine (Braunstein LZ *et al*. in press), the current study expands upon these findings to study branded “cool mint” ranitidine, and generic prescription ranitidine over a range of nitrite concentrations and incubation times. Since gastric factors vary widely between individuals, including sodium nitrite concentrations and gastric emptying times, our *in vitro* results are limited in scope but generally represent physiologically relevant conditions. Namely, we show an increase in NDMA levels over time at pH 2.5 for both medication types (generic prescription and branded “cool mint”) regardless of starting sodium nitrite concentrations. Moreover, the trajectory of NDMA yield suggests that further production of NDMA is likely beyond the final time point of 4 hours. This finding may merit particular consideration in light of evidence that ranitidine slows gastric emptying^25,26^, potentially facilitating prolonged reaction times in the gastric milieu and yet higher yield of NDMA. Although these and other laboratory findings demonstrate a link between ranitidine and NDMA, the implications for human carcinogenesis remain poorly understood.

While prior studies have not specifically focused on several cancers analyzed in our cohort, the association with breast cancer has been investigated in a case-control study observing that users of ranitidine (but not the other H_2_-blockers, famotidine or cimetidine) had a 2.4-fold higher odds of developing hormone receptor-positive invasive ductal carcinoma, as compared to never-users.^7^ Here, we do not have detailed outcome information to replicate these analyses, nor would we have the power to do so. Further work in a well-powered study will be important for elucidating these site-specific subgroup differences.

Of note, we deliberately excluded gastric and esophageal cancers from our analyses due to the potential for reverse causality and confounding by indication. Those with gastroesophageal reflux have both a higher likelihood of receiving an H_2_-blocker and of developing overt gastroesophageal malignancies, thereby complicating determinations of causality. In addition, secular trends in clinical prescribing patterns for peptic ulcer disease or reflux could obscure potential underlying associations between H_2_-blockers, PPIs and gastroesophageal cancers in our analysis within an oncology cohort.

There are several important strengths to this analysis. First, this paper addresses a question of public health relevance: many people have been exposed to this drug, and yet little is known about its association with cancer risk. Moreover, to address concerns regarding the potential for confounding by indication, active comparator analyses were conducted. Even so, results must be considered in the context of study limitations. These epidemiologic analyses were conducted entirely within an oncologic cohort and may not be generalizable to the broader public. Moreover, as with many such observational studies, exposure information was ascertained from self-reported histories with no available data regarding dose or duration of use. As a result, it remains unclear to what extent our findings apply to the spectrum of available dosing regimens or durations of ranitidine use. In addition, we were unable to evaluate the reasons for initiating a PPI or H_2_-blocker, and we have no data on prior use, such that reverse causality cannot be excluded. Despite comprising a large cohort, analyses of ranitidine and less-common cancers were not well-powered and remain exploratory in nature. Even among the more common cancer analyses, it is important to note that the entire cohort represents an oncologic population, such that odds ratios represent the odds of presenting with a certain cancer as compared to others, and not the odds of developing a certain cancer more generally in a typical population (because there were no patients in our cohort who were free of cancer). Whereas “controls” in similar studies are typically unaffected patients, in this analysis they are those with other malignancies (i.e. we found the odds of breast, thyroid, bladder and prostate cancers to be elevated in relation to other cancers, not in relation to being free of cancer). We were similarly unable to examine associations by subgroups of interest, and therefore, results may be obscured for specific subtypes. This is of particular relevance for cancers with known etiologic heterogeneity, such as low versus high grade prostate cancer, estrogen-receptor positive versus negative breast cancers and the various subtypes of thyroid cancer, etc. Indeed, all associations identified in this study should be viewed as hypothesis-generating.

Thus, we demonstrate that under a variety of physiologic reaction conditions, ranitidine yields elevated amounts of NDMA. Moreover, in a cohort of cancer patients taking H_2_-blockers or PPIs at the time of their cancer diagnosis, we found an association between ranitidine use and cancers of the breast, prostate, thyroid and bladder, as compared to other cancers. Unexplained inverse associations were observed with ranitidine and cancers of the brain and colorectum. Analyses of larger populations with longer follow-up may further elucidate important associations, particularly for less common malignancies.

## Data Availability

The patient-level data comprising the analyses in this study are HIPAA protected.
**The authors respectfully request that this preprint not be posted prior to January 29, 2021**

## Acknowledgement

LZB, KO and EK were supported by grant P30CA008748 from the National Cancer Institute.

AH, KK, QW and DYL are employees of Valisure, LLC, which conducted all chemical and molecular assays reported herein.

## Notes

### Competing Interest Statement

Amber Hudspeth MSc, Kaury Kucera PhD, Qian Wu PhD, David Y. Light BSc are employed by Valisure LLC.

### Funding Statement

LZB, KO and EK were supported by grant P30CA008748 from the National Cancer Institute. The authors received no other funding for any aspect of the submitted work.

### Author Declarations

The institutional review board of Memorial Sloan Kettering Cancer Center approved this the conduct of this study.

